# Vaccine beliefs, adverse effects, and quality of life in patients with cancer undergoing routine COVID-19 vaccination

**DOI:** 10.1101/2024.06.02.24308345

**Authors:** Amy Body, Mark Donoghoe, Luxi Lal, Claire Wakefield, Elizabeth Stephanie Ahern, Antoinette Anazodo, Noemi Auxiliadora Fuentes Bolanos, Bhavna Padhye, Peter Downie, Stephen Opat, Nada Hamad, Michael Francis Leahy, Vivienne Milch, Eva Segelov

## Abstract

**Background:** Concerns about side effects and treatment interactions and delays may contribute to COVID-19 vaccine hesitancy amongst cancer patients. In the large prospective SerOzNET study of COVID-19 vaccine response in children and adults with cancer, vaccine beliefs, physician- and participant-reported adverse events (AE), treatment interruptions and quality of life (QoL) were studied.

**Methods:** The Australian experience with COVID-19 gave a unique opportunity to study vaccination response in an infection- and vaccine-naïve cancer population. Patients with current or recent solid or hematological malignancy, aged five and over, had serial assessments prior to and following multiple SARS-CoV-2 vaccinations. Electronic surveys were administered at baseline and after first, second and third doses to collect vaccine beliefs (Oxford Confidence and Complacency Scale), patient-reported toxicity and QoL (QLQC30 or PedsQL). Detailed toxicity data were collected at clinic visits and from medical records.

**Results:** A total of 1385 vaccination doses were administered (93% BNT162b2), with at least 1 dose received by 499 patients, of whom only seven had known prior COVID-19 infection. Vaccine related beliefs were generally positive. There were no vaccine-related interruptions to cancer therapy. AE occurred in 95% of recipients, with the highest ranked severity being mild in 36% and moderate, severe or serious in 31%, 19% and 6% respectively. QoL showed no significant deterioration post-vaccination.

**Conclusion:** This robust dataset provides evidence regarding safety and tolerance of SARS-CoV-2 vaccination in adults and children with cancer. Patients and families can be reassured that rates of AEs are comparable to the general population and do not impact delivery of cancer therapy or QoL.

## 1. Background

The COVID-19 pandemic was highly disruptive to cancer care, with significant morbidity and mortality amongst medically vulnerable people^1^. COVID-19 remains a concern for patients with cancer, who are advised to receive booster vaccinations^2^. Some patients exhibit vaccine hesitancy, strongly influenced by vaccine-related beliefs^3^. Concerns about toxicities, impact on treatment schedules and interactions with cancer therapy also contribute. There is little data on which to base informed shared decision-making about benefits of COVID-19 vaccination^4^.

Data demonstrating the safety of COVID-19 vaccination in patients with cancer comes from limited clinical trials, where toxicities appear similar to the general population^5^. However, many patients had prior COVID-19 exposure and most data were collected during the pandemic peak^6^. There is minimal information on interruptions to cancer care and to our knowledge, no reports regarding effects of vaccination on participant-reported quality of life (QoL)^7^ ^8^.

Due to early lockdown measures including border closures, Australia in 2021 had very low prevalence of COVID-19 infection^9^. The SerOzNET study was initiated in this setting, enrolling vaccine-naïve adults and children with cancer. Common data elements from the National Cancer Institute’s Serological Sciences Network for COVID-19 (SeroNet) framework were collected^10^, including secondary endpoints of vaccine beliefs, toxicity data (physician reported and Patient Reported Outcome Measures -PROMS), interruptions to cancer treatment and QoL^11^ ^12^. This study is highly relevant to future care of cancer patients, who will decide with their clinicians about ongoing COVID-19 protective measures.

## 2. Methods

SerOzNET (ACTRN12621001004853) is a prospective study of patients with cancer undergoing SARS-CoV-2 vaccination in Australia, involving serial blood collection for assessment of immune response (primary endpoint) and collection of outcomes related to participant experiences (secondary endpoint and focus of this paper). All participants or guardians provided written informed consent. The protocol is published and approved by Monash Health Human Research Ethics Committee (RES-21-0000-337A)^13^.

### Study Setting

Enrolment occurred from June 2021 to December 2022 at five centers across Australia, including three specialist childhood cancer centers. The lead site is a culturally diverse outer metropolitan health service with a catchment area of 1.8 million people, one third of whom were born outside Australia.

### Study population

Participants were ≥ 5 years and in one or more of the following clinical groups: receiving systemic anti-cancer therapies (chemotherapy, targeted or hormonal therapy, immunotherapy); completed cytotoxic chemotherapy within 12 months; current hematological cancer associated with immune compromise regardless of treatment status. At enrolment, all patients were COVID vaccine naïve. Although not an exclusion criterion, most (>98%) had not had COVID-19 infection. Exclusion criteria were life expectancy < 12 months; inability for regular venipuncture and pregnancy.

Patients received vaccinations according to government regulations which were frequently updated during the study period (Figure 1)^14^. Briefly, patients ≥ 60 years were eligible for mRNA vaccine (BNT162b2 or mRNA-1273) or viral vector vaccine (ChAdOx1-S), until the latter was discontinued due to reports of vaccine-induced thrombotic thrombocytopenia. Patients < 60 years were only offered mRNA vaccines.

**Figure 1:**
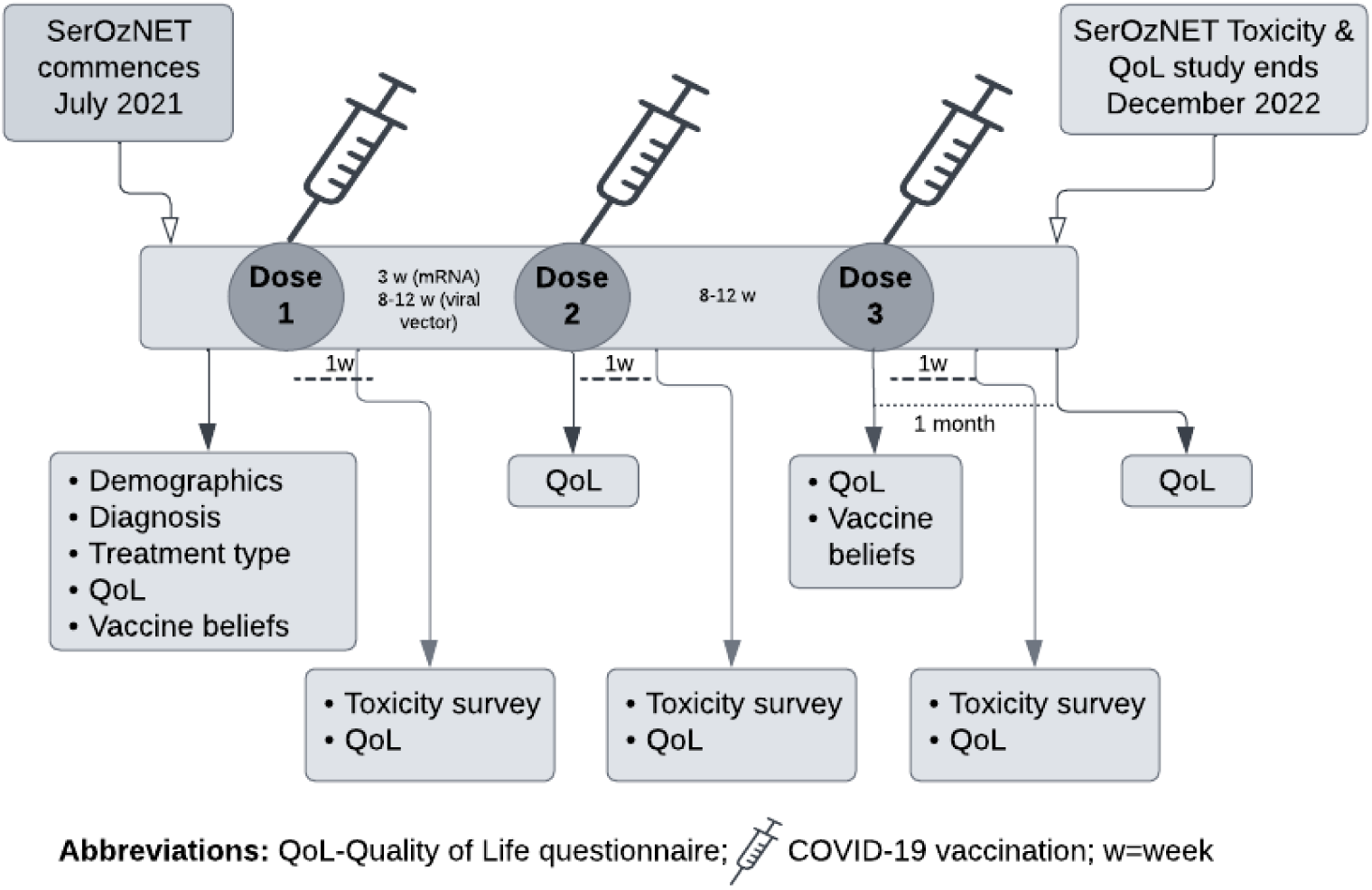
Study timeline

### Data collection and storage

Data were managed using REDCap® (Research Electronic Data Capture) hosted by Monash University^15^ ^16^.

### Demographics, time points and outcomes

Age, sex, ethnicity, country of birth, cancer type, stage and treatment type were collected at baseline.

Vaccine-related beliefs were assessed at baseline using the Oxford COVID-19 Vaccine Confidence and Complacency Scale (OCCS), where higher scores indicated increased level of concern, up to maximum score of 100^3^. A modified version (removing 2 items regarding speed of vaccine development that were not relevant after initial dose) was completed by adults prior to 3^rd^ vaccination dose (Appendix 1A).

Safety data were extracted from electronic medical records (Appendix 1B). Two time periods were assessed: from enrolment until 30 days post-2nd vaccination (hereafter called the initial two-vaccine period) and from prior to 3^rd^ vaccination until 30 days later (hereafter called the first booster period). Causality of AEs was categorized by investigators (hematologists/oncologists) as: related, possibly related, or not related to vaccine. Grading used Common Terminology Criteria for Adverse Events version 5.0.^17^.

PROMs were collected by surveys sent to participants’ phones, or electronic tablets on site. Non-English-speaking patients completed surveys with a telephone interpreter. For children > 7 years, surveys for both child and parent were collected; < 7 years, surveys were sent to parents only.

Participant-reported toxicity was collected 7 days after each of the first three vaccine doses, using relevant Patient-Reported Outcome Common Terminology for Adverse Events items, plus five questions on treatment interruptions and healthcare utilization (Appendix 1C)^18^.

QoL was assessed using QLQ-C30 for adults and age-appropriate PedsQL cancer modules (child and/or parent report forms) administered prior to first and second vaccinations, one week after each, and one month after final dose^19^ ^20^.

### Statistical analysis

The full statistical analysis plan is available (Appendix 2).

OCCS scores were summarized descriptively. The Spearman correlation coefficient [and 95% confidence interval (CI)] was calculated between OCCS total score and QoL subscales at baseline. Patient- and physician-reported toxicities were summarized descriptively. A binary variable was also calculated, to capture participant-reported toxicities after dose 1 or 2, as well as the worst severity of any toxicity. Likelihood ratio tests were used to probe association between worst participant-reported toxicity and the probability of receiving dose 3 (logistic regression), comparing incidence of each physician-reported toxicity (logistic regression), incidence rate of each physician-reported toxicity (Poisson regression accounting for follow-up time via an offset), and incidence and incidence rate of physician-reported serious adverse events (SAEs) (Poisson regression with and without a follow-up time offset), comparing patients who received different vaccine types.

For analyses with > 5% missing data, or evidence that data absence was not completely random, multiple imputation was used to ensure results were valid under a missing-at-random assumption. Testing was performed for association between dose 3 OCCS score and worst participant-reported toxicity at doses 1-2, adjusting for baseline OCCS score by using ANCOVA within each imputed dataset, and combining across imputations using the D2 statistic^21^. A flexible association between participant-reported toxicity incidence and selected QoL scores was allowed by fitting logit-link binomial generalized additive models to each imputed dataset, taking the median p-value from the approximate test of no association as an overall test of significance^22^. The association between participant-reported AEs and baseline QoL was assessed for adult patients only, due to small numbers of children.

## 3. Results

Of 511 consented participants, 499 received at least one vaccination dose and form the cohort for this analysis. Characteristics of the 107 children (5-19 years) and 392 adults are shown in Table 1.

**Table 1:** Baseline demographics and clinical characteristics.

| **Demographics** | **Adults(n=392)** | **Children (5-19y)(n=107)** | **Overall(n=499)** |
| --- | --- | --- | --- |
| **Age (y)** |  |  |  |
| Mean (SD) | 56.5 (14.1) | 12.4 (3.8) | 47.1 (22.1) |
| Median (IQR) | 58.5 (47.0, 66.0) | 13.0 (9.5, 15.0) | 52.0 (29.5, 64.0) |
| Range | 20, 85 | 5, 19 | 5, 85 |
| **Sex (%)** |  |  |  |
| Male | 160 (41) | 66 (62) | 226 (45) |
| Female | 232 (59) | 40 (37) | 272 (55) |
| Other | 0 | 1 (1) | 1 |
| **Household language (%)** |  |  |  |
| English | 292 (75) | 93 (89) | 385 (78) |
| Other | 99 (25) | 12 (11) | 111 (22) |
| Unknown | 1 | 2 (2) | 3 |
| **Country of birth (%)** |  |  |  |
| Australia | 197 (50) | 92 (88) | 289 (58) |
| Other | 194 (50) | 13 (12) | 207 (42) |
| Unknown | 1 | 2 (2) | 3 |
| **Indigenous status (%)** |  |  |  |
| Aboriginal and/or Torres Strait Islander | 3 (1) | 4 (4) | 7 (1) |
| **Cancer diagnosis (%)** |  |  |  |
| Head and neck | 12 (3) | 1 (1) | 13 (3) |
| Gastrointestinal | 59 (15) | 2 (2) | 61 (12) |
| Thoracic | 18 (5) | 0 (0) | 18 (4) |
| Bone, connective tissue | 1 (0) | 18 (17) | 18 (4) |
| Skin | 5 (1) | 0 (0) | 5 (1) |
| Breast | 82 (21) | 0 (0) | 82 (16) |
| Gynecological | 33 (8) | 0 (0) | 33 (7) |
| Genitourinary | 31 (8) | 2 (2) | 31 (6) |
| Central nervous system | 2 (1) | 7 (7) | 7 (2) |
| Lymphoma | 74 (19) | 19 (18) | 93 (19) |
| Myeloma | 25 (6) | 0 (0) | 25 (5) |
| Leukemia | 36 (9) | 49 (46) | 85 (17) |
| Other | 13 (3) | 9 (8) | 22 (4) |
| Unknown/Missing | 1 (0) | 0 (0) | 1 (0) |
| **Treatment type\*(%)** |  |  |  |
| Chemotherapy | 165 (42) | 94 (88) | 259 (52) |
| Immunotherapy | 59 (15) | 5 (5) | 64 (13) |
| Hormone therapy | 57 (15) | 0 (0) | 57 (11) |
| Bone modifying agent | 10 (3) | 0 (1) | 11 (2) |
| Targeted therapy only | 49 (12) | 3 (3) | 52 (10) |
| Combined chemo-immunotherapy | 42 (11) | 7 (7) | 49 (10) |
| No systemic therapy | 25 (6) | 1 (1) | 26 (5) |
| Other | 19 (5) | 2 (2) | 21 (4) |

\*May add to >100% as some patients were on multiple treatments; y=years

### 3.1. Cohort diversity and engagement with surveys

There were 49 primary languages, with 12% of children and 50% of adults born outside Australia. Survey completion rates after doses 1 and 2 were > 90% for adults, regardless of country of birth or language (Appendix 3A). For children, response ranged from 27-57%, with < 10% difference according to place of birth of child or parent.

### 3.2. Vaccine-related beliefs

Overall, vaccine-related beliefs had positive themes. For adults, 30% believed they would possibly, probably, or definitely contract COVID-19 in the next 12 months; 59% believed the vaccine would either probably or definitely work for them and 89% believed that getting the COVID-19 vaccine would be helpful or really helpful for the community around them. Only 11% believed the vaccine would be moderately unpleasant or painful.

Of the 57 children who responded, 49% believed they would possibly, probably, or definitely contract COVID-19 in the next 12 months; 63% believed the COVID-19 vaccine would probably or definitely work for them; 84% believed getting the vaccine would be helpful or really helpful for the community around them and only 16% believed the vaccine would be moderately unpleasant or painful.

The median total OCCS at baseline was 23.8, with adjusted median score (removing questions regarding speed of vaccine development) of 23.9; and pre-third dose score 22.8 (Appendix 3B). There was no evidence that the latter was associated with the severity of participant-reported AE, after adjusting for baseline score (F-test p=0.42). The median OCCS score was similar between children and their parents (Appendix 3c).

### 3.3. Investigator-reported adverse events

#### 3.3.1. Delays to cancer treatment

During the initial two-vaccine period (mean of 56 days; 53 and 89 days for mRNA and viral vaccines respectively), treatment delays occurred in 9% of adults and 20% of children and treatment modifications were recorded in 17% and 14% respectively. (Table 2). In the first booster period, therapy delays were reported in 5% of adults and 9% of children and regimen modifications in 8% and 9% respectively. No delays or dose modifications were attributed to the vaccine.

**Table 2:** Physician reported modifications and delays to treatment.

|  | Adults |  | Children |  |
| --- | --- | --- | --- | --- |
|  | Doses 1-2 | Dose 3 | Doses 1-2 | Dose 3 |
| **Cancer treatment delay (% , 95% CI)** |  |  |  |  |
| Nil | 346 (91, 87-93) | 292 (95, 92-97) | 83 (81, 72-88) | 59 (91, 81-97) |
| Vaccine related | 0 (0 0-1) | 0 (0, 0-1) | 0 (0, 0-4) | 0 (0, 0-6) |
| Cancer treatment toxicity | 23 (6 4-9) | 8 (3, 1-5) | 11 (11, 5-18) | 4 (6, 2-15) |
| Other cause* | 13 (3, 2-6) | 8 (3, 1-5) | 9 (9, 4-16) | 2 (3, 0-11) |
| **Cancer treatment modification (% , 95% CI)** |  |  |  |  |
| Nil | 316 (83, 79-87) | 283 (92, 88-95) | 87 (86, 78-92) | 59 (91, 81-97) |
| Vaccine related issue | 0 (0, 0-1) | 0 (0, 0-1) | 0 (0, 0-4) | 0 (0, 0-6) |
| Sompleted planned treatment | 15 (4, 2-6) | 5 (2, 1-4) | 2 (2, 0-7) | 3 (5, 1-13) |
| Treatment AE/ intolerance | 21 (6, 3-8) | 9 (3, 1-5) | 10 (10, 5-17) | 0 (0, 0-6) |
| Disease progression | 23 (6, 4-9) | 11 (4, 2-6) | 0 (0, 0-4) | 2 (3, 0-11) |
| Other** | 6 (2, 1-3) | 0 (0, 0-1) | 2 (2, 0-7) | 1 (2, 0-8) |

\*Other reasons for cancer treatment delays included: **Adults** Dose 1-2; cancer related issue, 7, intercurrent illness, 4, patient preference, 2. Dose 3; intercurrent illness, 5, patient preference, 3. **Children** Dose 1-2; cancer related issue, 2, intercurrent illness, 6, patient preference, 1. Dose 3; intercurrent illness, 2.
**Other reasons for treatment modifications included:** Adults **Dose 1-2; treatment optimization, 4, intercurrent illness, 1, patient decision, 1.** Children **\*\*Dose 1-2; cancer related issue, 1, infection, 1. Dose 3; treatment optimization, 1.**

#### 3.3.2. Serious Adverse Events

Grade and attribution of AEs are detailed in Appendix 3D. Rates of SAE during the initial two-vaccine period in adults and children were 12% and 30% respectively; during the first booster period were 3% and 19%. Most SAEs were deemed not related to the vaccine. During the initial period, 9% SAE in adults and 45% in children were deemed possibly related; none were reported as definitely related. During the first booster period, 0 and 25% of SAE in adults and children respectively were possibly related (Appendix 3E). Only three SAEs in children (two fever, one headache) were deemed “definitely or very likely” related to vaccine.

#### 3.3.3. Allergic and thrombotic events

No allergic reactions were reported. In adults, there was one deep vein thrombosis, one pulmonary embolism, two other venous thrombosis and one ischemic stroke. Each occurred during the initial two-vaccine period. No thrombotic events were reported for children.

#### 3.3.4. Other adverse events

Amongst adults, grade 1 toxicities (fever, rash, headache, liver function abnormalities on chemotherapy) were reported in one instance each; fatigue was reported twice. One adult had grade 2 chest pain determined to be non-cardiac. Amongst children, several grade 1 potentially attributable toxicities (headache, dizziness, fatigue, malaise, fever, nausea, vitiligo, abdominal pain, chest pain with normal investigations) were reported in one instance each; vomiting was documented in two patients; one adolescent had grade 4 thrombocytopenia on radiotherapy.

Lymphadenopathy after vaccination was reported in 29 adults (8%) in the initial two-dose period and 11 (3%) in the first booster period; only four cases were attributed to vaccine. Two children had lymphadenopathy reported, neither requiring intervention.

### 3.4. Participant-reported adverse events

At least one AE was reported by 85%, 88% and 86% of adults after the first three doses respectively; for children this was 94%, 88% and 93%. The majority were mild to moderate severity (Figure 2 and Figure 3). In adults, self-reported “severe” or “serious” AEs after the first three doses occurred in 13%, 26% and 25% respectively. Fatigue was most frequent, reported 70 times. In children, self-reported “severe” or “serious” AEs occurred in 6%, 11% and 22% after each dose respectively. The most common was fever, reported in six instances.

**Figure 2:**
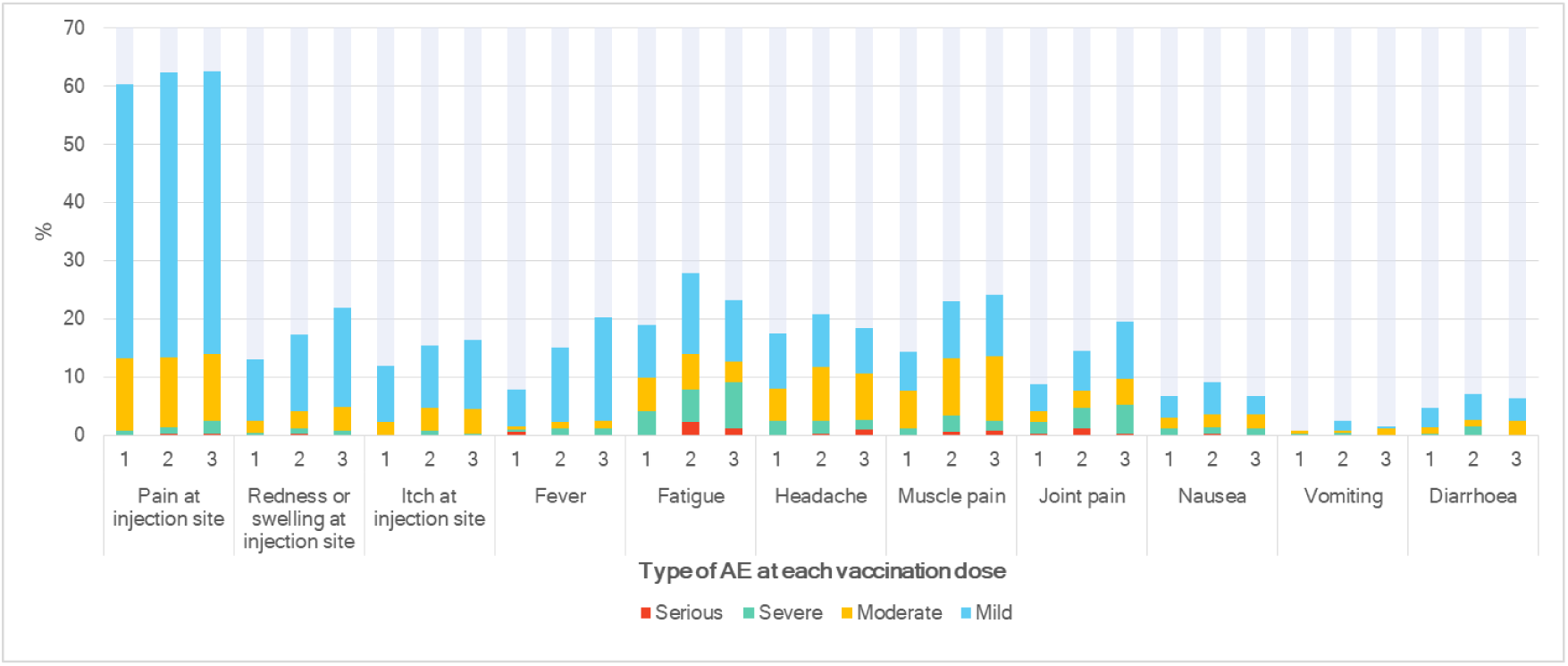
Patient reported adverse event frequency - Adults. Grade according to Patient-Reported Outcomes Common Terminology Criteria for Adverse Events^18^

**Figure 3:**
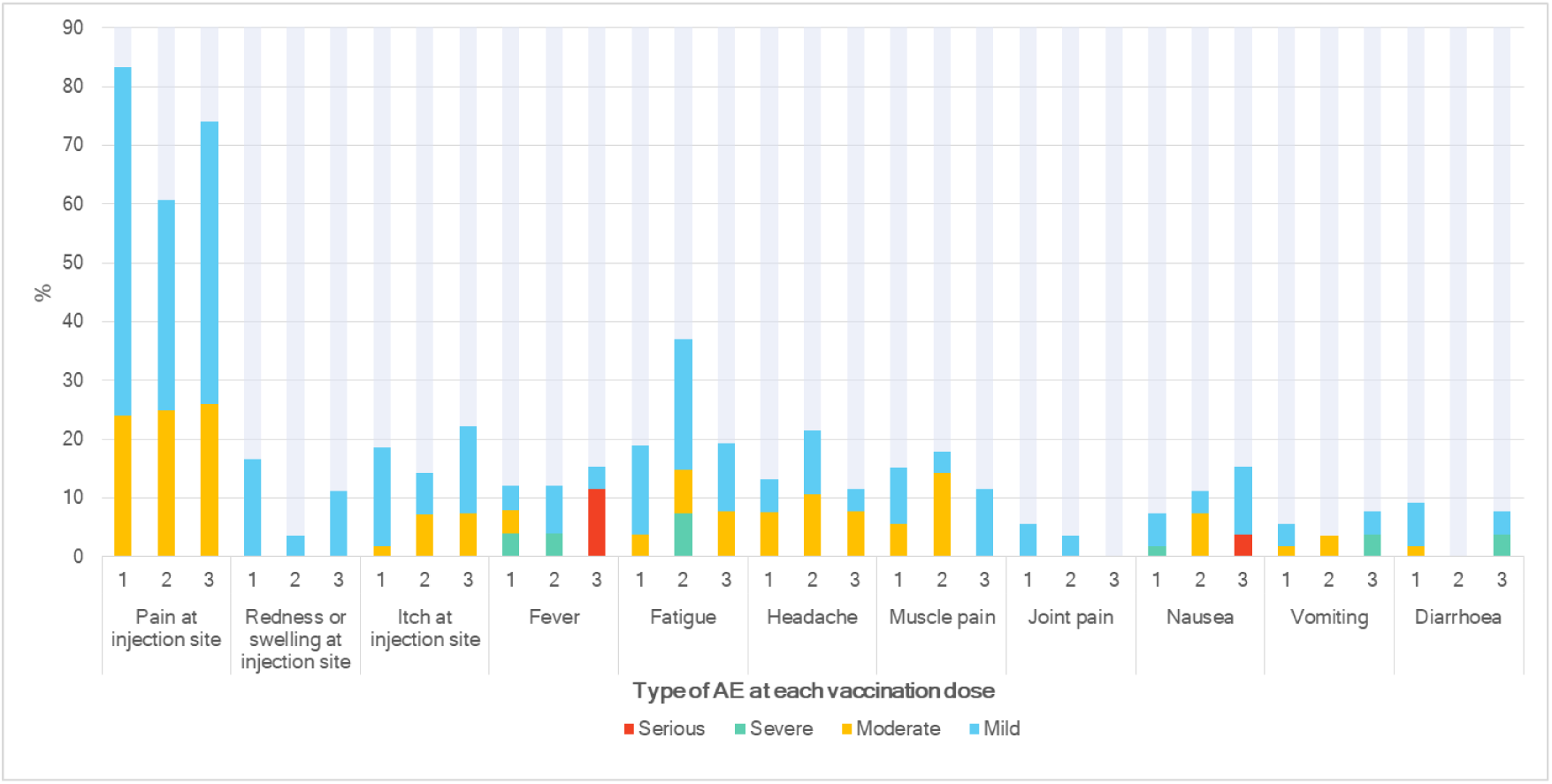
Patient reported adverse event frequency - Children. Grade according to Patient-Reported Outcomes Common Terminology Criteria for Adverse Events^18^

Local side effects occurred in 65-85% of participants, depending on age and dose, and were more common than systemic side effects, in 50-62% (Appendix 3F, 3G). Healthcare utilization (visiting doctor or emergency department) was reported in 6-10% adults and 8-31% of children. Participant-reported interruptions or delays to cancer treatment for any reason were infrequent (0-7%).

The severity of participant-reported AE after the first two vaccinations was not significantly associated with receipt of a third dose (Appendix 3H).

#### 3.4.1. Adverse events by vaccine type

Comparing events by vaccine type was only possible in patients over 60 years. As only two patients in this group received a viral vector vaccine for the third dose, most comparisons examine outcomes post dose 1 and 2 (Appendix 3I). There was no statistical difference in investigator-reported AE between mRNA and viral vector vaccines, however, events were rare (n= 145). SAE were more common in the viral vector group however, after adjusting for time between vaccine doses, the difference was non-significant (RR 1.81, 95% CI 0.76-4.20) (Appendix 3J). Participant-reported injection site pain was higher with mRNA compared to viral vector vaccines; all other participant-reported AEs occurred at similar rates (Appendix 3K).

### 3.5. Quality of life

Adult QoL summary scores were stable pre and post doses 1 and 2. A sensitivity analysis conducted with multiple imputation for missing data did not alter these findings (Appendix 3L). QLQC30 functional and symptom scales showed no differences before and after doses for all scales except pain, which was slightly increased.

Self-reported PedsQL showed small improvements in several domains after doses 1 and 2 (Appendix 3M). Parent-report PedsQL functional and symptom scales showed no difference pre- and post-doses, except for slight reduction in pain post dose 1.

Lower QoL scores were associated with more negative vaccine beliefs. In adults, there was a small but significant negative correlation seen for emotional functioning, global health status and the vaccine-related beliefs summary score. For children, correlation coefficients were negative between five baseline QoL measures and vaccine-related beliefs, but with high uncertainty due to small sample size (Appendix 3N).

## 4. Discussion

SerOzNET is a comprehensive, prospective study of outcomes reported by children and adults with cancer and their physicians regarding vaccine beliefs, toxicity, QoL, and impact on cancer treatment of COVID-19 vaccination. There was low major toxicity, mild toxicity as expected, no deterioration in QoL after vaccination, and minimal treatment interruptions.

Vaccine beliefs were largely positive, consistent with participants who agreed to vaccination. We confirmed the hypothesis that a lower baseline QoL predicted more negative vaccine beliefs. This identifies an opportunity in designing vaccination strategies for vulnerable populations, to optimize QoL which might encourage positive attitudes towards preventative interventions.

Concerns about delaying vaccination or interference with therapy schedules is reported as a driver of hesitancy in the cancer population^4^ ^23^. Our finding of no delays attributable to vaccination should help allay concerns, supporting reports from smaller cohorts^7^ ^8^.

Confirming comparable rates of AE in this large, real-world population to the general population is also significant. Local and systemic adverse effects reported through PROMS were rarely documented in medical records, thus not included in investigator-reported toxicity, highlighting the importance of patient reporting. Conversely, some hospital admissions not reported by participants were recorded on physician review. This study highlights that patient and physician reports are complementary and should be used together for assessment of safety.

Comparison of SAE rates from SerOzNET with similar studies is limited by different reporting periods. The large “VOICE” study (patients with solid cancers in The Netherlands receiving mRNA vaccines) reported that 25% of SAE were potentially linked to vaccine, and that vaccine-attributable SAE were generally reversible^24^. Rates of venous thromboembolic events were similar, with arterial events rare. These concordant findings are reassuring, particularly with background incidence of thrombosis documented around 12.6% per year for ambulatory patients on chemotherapy^25^ ^26^.

Overall, quality of life was stable pre- and post-vaccination. Change in pain scores was discordant between adults (pain increased) and children (reduced), but the two point difference in adults is less than the minimal clinically important difference of at least 4 points^27^. Hence, the finding is likely incidental; it may also reflect the highly dynamic nature of pain.

The SerOzNET study invested significant effort in engagement of non-English speaking and overseas-born participants, as these groups are commonly less well represented, if not excluded from QoL and PROMs studies. Use of telephone interpreters allowed non-English speaking patients to be included, evidenced by findings of similar rates of survey completion throughout the study. Previous data have shown bias toward patients of white ethnicity being included in psychosocial studies of oncology patients^28^. A potential source of bias with this approach is that the English-speaking patients completed their PROMs in private, without direct involvement of study staff, and that not all surveys were validated when translated, although where available, official translations were used. This complexity has been noted by other investigators, however, no consensus has been reached^29^. Overall, we found the pragmatic use of telephone interpretation for PROMs was acceptable to patients and allowed inclusion of English-language PROM tools, important in a setting where a very large number of languages spoken makes written translation of materials less feasible.

In the childhood cohort, survey response rates were lower, likely reflecting care demands. Prior studies noted variable engagement with PROMs in the paediatric setting, with lower completion of electronic self-reports in the outpatient setting^28^ ^30^^31^. Our study was limited to delivery of surveys to a single phone number per family. Resources permitting, incorporation of PROMs at clinic visits rather than remote electronic delivery may result in higher levels of pediatric engagement.

Major strengths of this study are the number and diversity of patients enrolled, and the unique COVID-19 environment in which recruitment occurred, capturing individuals who were both COVID-19- and vaccine-naïve^32^. Furthermore, detailed PROMs, including QoL post vaccination and vaccine beliefs, have not been systematically assessed in cancer populations post COVID-19 vaccination.

### 4.1 Limitations

SerOzNET was undertaken during recurrent lockdown periods, with frequent changes in public health vaccine policy. These gave rise to various methodological and practical limitations, restricting the number of requests without fatiguing patient engagement. SerOzNET incorporated formal Patient and Public Involvement from initial study design and throughout its conduct.

Inclusion of a patient representative on the Study Management Committee allowed appreciation of survey fatigue as vaccination extended beyond the initial two dose schedule. Ultimately, this led to discontinuation of surveys for later doses, which may have provided additional longitudinal information. Finally, SerOzNET was undertaken in a real-world setting, with demands on participants’ and carers time, within a tight budget. Under these constraints, engagement of children in particular was a challenge; nevertheless, to our knowledge this is the largest childhood cancer COVID-19 vaccination study reported.

### 4.2 Conclusion

This detailed, extensive real-world study provides evidence that SARS-CoV-2 vaccination for adults and children with cancer is tolerable and does not result in significant disruption to cancer therapy or deterioration in quality of life. Patients with cancer can be reassured that SARS-CoV-2 vaccination is safe during and after cancer treatment. The ability to quote these data in discussions with patients and families contributes to the evidence base for shared decision making in this critical area.

#### Inclusion and Diversity

We support inclusive, diverse, and equitable conduct of research.

## Data Availability

All data produced in the present study are available upon reasonable request to the authors.

## Acknowledgments

The SerOzNET study team would like to thank all of our study participants. We thank Cancer Australia for funding support and oversight throughout the course of the study, and our additional funders The Victorian Cancer Agency, The Leukaemia Foundation (Australia), and The Cancer Network of Western Australia. We also thank our study collaborators and co-investigators.

The funding institutions did not dictate the design of the study, have access to the data, or influence the decision to submit the manuscript for publication.

## Disclaimers

None.

## Data sharing statement

The study data can be accessed by contacting the corresponding author.

