## Appendix 1 for "Vaccine beliefs, adverse effects, and quality of life in patients with cancer undergoing routine COVID-19 vaccination"

### Appendix 1- Methods

**Appendix 1a Modified Oxford COVID-19 Vaccine Confidence and Complacency Scale (OCCS) for dose 3**

| | |
|---|---|
| Section Header: *Your opinions about the additional (booster) dose of the COVID-19 vaccine*<br>Do you think you will be infected with COVID-19 over the next 12 months? | radio<br>1 Definitely<br>2 Probably<br>3 Possibly<br>4 Probably not<br>5 Definitely not<br>6 Don't know |
| The additional dose (booster) COVID-19 vaccine is likely to: | radio<br>1 Work for almost everyone<br>2 Work for most people<br>3 I am unsure how many people it will work for<br>4 Not work for most people<br>5 Not work for anyone<br>6 Don't know |
| The additional dose (booster) COVID-19 vaccine is likely to: | radio<br>1 Definitely work for me<br>2 Probably work for me<br>3 May or may not work for me<br>4 Probably not work for me<br>5 Definitely not work for me<br>6 Don't know |
| If I get the additional dose (booster) COVID-19 vaccine it will be: | radio<br>1 Really helpful for the community around me<br>2 Helpful for the community around me<br>3 Neither helpful nor unhelpful for the community around me<br>4 Unhelpful for the community around me<br>5 Really unhelpful for the community around me |

| | |
|---|---|
| | 6 Don't know |
| If individuals like me get the additional dose (booster) COVID-19 vaccine it will: | radio<br>1 Save a large number of lives<br>2 Save some lives<br>3 Have no impact<br>4 Lead to more deaths<br>5 Lead to a large number of deaths<br>6 Don't know |
| If many people do not get the additional dose (booster) vaccine this: | radio<br>1 Will be dangerous<br>2 May be dangerous<br>3 Will have no consequences at all<br>4 May be good<br>5 Will be good<br>6 Don't know |
| I expect that receiving the vaccine will be: | radio<br>1 Hardly noticeable<br>2 A little unpleasant<br>3 Moderately unpleasant<br>4 Painful<br>5 Extremely painful<br>6 Don't know |
| The side effects of getting the additional dose (booster) COVID-19 vaccine will be: | radio<br>1 None<br>2 Mild<br>3 Moderate<br>4 Significant<br>5 Life threatening<br>6 Don't know |
| The additional dose (booster) COVID-19 vaccine will: | radio<br>1 Greatly strengthen my immune system<br>2 Strengthen my immune system<br>3 It will neither strengthen nor weaken my immune system<br>4 Weaken my immune system<br>5 Greatly weaken my immune system<br>6 Don't know |

| | |
|---|---|
| Taking the additional dose (booster) COVID-19 vaccine: | radio<br>1 Will give me added confidence to get on with life just as before<br>2 Will give me greater freedom<br>3 Will have no effect on my confidence or freedom<br>4 Will restrict my freedom<br>5 Will completely restrict my freedom to get on with life<br>6 Don't know |
| Taking the additional dose (booster) COVID-19 vaccine will make me feel like a guinea pig | radio<br>1 Do not agree<br>2 Agree a little<br>3 Agree moderately<br>4 Agree a lot<br>5 Completely agree<br>6 Don't know |
| My experience with the previous vaccine doses makes me worried about receiving an additional dose | radio<br>1 A lot<br>2 Quite a bit<br>3 A little bit<br>4 Not at all<br>5 I'm not sure either way |
| I believe that an additional vaccine dose is a good idea for me and other people with similar medical conditions to me | radio<br>1 Strongly agree<br>2 Agree<br>3 Neither agree nor disagree<br>4 Disagree<br>5 Strongly disagree<br>6 Don't know |
| Regarding the timing of the extra dose- I think this is: | radio<br>1 Too early<br>2 About right<br>3 Too late<br>4 I am unsure either way<br>5 I don't think it is necessary |

**Appendix 1b Medically ascertained adverse event case report form**

a. Was there a serious adverse event (SAE) dose 1 until 1 month post dose 2?
b. Was there an SAE from dose 3 until 1 month post dose 3?
c. For each SAE: Date- SAE term as per CTCAE- Was the SAE attributable to the vaccine according to the treating physician?
d. Was cancer treatment delayed from any time between dose 1 and one month post dose 2?
   Yes- vaccine related
   Yes- treatment toxicity
   Yes- Other
   No
e. Reason for vaccine-related dose delay
f. Was cancer treatment delayed from any time between dose 3 and one month post dose 3?
   Yes- vaccine related
   Yes- treatment toxicity
   Yes- Other
   No
g. Reason for vaccine-related dose delay
h. Cancer treatment modification from dose 1 until 1 month post dose 2?

| | |
|---|---|
| 1 | Yes- completed planned treatment |
| 2 | Yes- intolerance to one of treatment drugs or adverse event related to treatment drug |
| 3 | Yes- disease progression |
| 4 | Yes - other- insert reason |
| 5 | Yes- vaccine related issue |
| 0 | No |

i. Cancer treatment modification from dose 3 until 1 month post dose 3?

| |
|---|
| Yes- completed planned treatment |
| Yes- intolerance to one of treatment drugs or adverse event related to treatment drug |
| Yes- disease progression |
| Yes - other- insert reason |
| Yes- vaccine related issue |
| No |

j. Thrombotic event between dose 1 and one month post dose 2?

| | |
|---|---|
| 0 | No |
| 1 | Yes- deep vein thrombosis |
| 2 | Yes - pulmonary embolism |
| 3 | Yes- other venous thrombosis (state) |
| 4 | Yes - coronary artery event |
| 5 | Yes- cerebrovascular event |
| 6 | Yes- other arterial thrombus (state |

k. Thrombotic event between dose 3 and one month post dose 3?
l. Allergic reaction due to dose 1- 2 or 3?
m. Decline in performance status between dose 1 and one month post dose 2
n. Decline in performance status between dose 3 and one month post dose 3?
o. Lymphadenopathy seen clinically/on imaging between dose 1 and one month post dose 2
p. Lymphadenopathy seen clinically/on imaging between dose 3 and one month post dose 3
q. Any other AE potentially attributable to vaccine during study period? Define as per CTCAE (term and grade)

**Appendix 1c Patient-Reported Outcomes version of the Common Terminology Criteria for Adverse Events (PRO-CTCAE®) questions**

| | |
|---|---|
| *Section 1 Side effect survey*<br>Did you have pain at the vaccine injection site (where the vaccine was injected into your arm)? | dropdown- Required<br>1 No<br>2 Mild- did not interfere with doing my usual activities<br>3 Moderate- slowed me down or made my usual activities more difficult<br>4 Severe- stopped me doing my usual activities<br>5 Serious- I needed to attend the emergency department due to pain at the injection site |
| Did you have redness or swelling at the injection site? | dropdown<br>1 No<br>2 Minor- 2 to 5cm area of redness or swelling<br>3 Moderate- 5 to 10cm area of redness or swelling<br>4 Severe- more than 10cm (most of upper arm affected)<br>5 Serious- blistering- skin damage or soft tissue damage requiring medical attention |
| Did you have itch at the injection site? | dropdown- Required<br>1 None<br>2 Mild- barely noticed it<br>3 Moderate- for example needed to scratch or use cool pack<br>4 Severe- needed to take medication<br>5 Very severe- needed to seek medical attention |
| Section Header: *Section 2*<br>Have you had a fever (a temperature) after the vaccination? | dropdown<br>1 No<br>2 Yes- I felt feverish but did not take temperature<br>3 Yes- mild 37.5-37.9 degrees<br>4 Yes- moderate 38-38.5 degrees<br>5 Yes- high fever 38.6 degrees or higher<br>6 Yes- went to hospital for assessment or treatment |
| Did you take any medication to treat side effects after your vaccine? | checkbox<br>0 medication___0 No<br>1 medication___1 Yes- paracetamol<br>2 medication___2 Yes - anti inflammatory for example ibuprofen (Nurofen)- diclofenac (Voltaren)- naproxen (Naprosyn) |

| | |
|---|---|
| | 3 \| medication___3 \| Yes- opioid pain killer - oxycodone (Endone)- codeine (Panadeine or Panadeine Forte)<br>4 \| medication___4 \| Yes- other- specify |
| What medication did you take to treat side effects after your vaccine? *Please write the name of the medication you used* | text (alpha_only) |
| Did you have new or worsened fatigue since your vaccination? | Yes/No- Required<br>1 \| Yes<br>0 \| No |
| If yes- In the last 7 days- how much did FATIGUE- TIREDNESS- OR LACK OF ENERGY INTERFERE with your usual or daily activities? | dropdown<br>1 \| Not at all<br>2 \| A little bit<br>3 \| Somewhat<br>4 \| Quite a bit<br>5 \| Very much |
| Did you have new or worsened headache after your vaccination? | Yes/No- Required<br>1 \| Yes<br>0 \| No |
| If yes- In the last 7 days- what was the SEVERITY of your HEADACHE at its WORST? | dropdown<br>1 \| None<br>2 \| Mild<br>3 \| Moderate<br>4 \| Severe<br>5 \| Very severe |
| In the last 7 days- how OFTEN did you have SHIVERING OR SHAKING CHILLS? | dropdown<br>1 \| Never<br>2 \| Rarely<br>3 \| Occasionally<br>4 \| Frequently<br>5 \| Almost constantly |
| Did you have new or worsened muscle pain since your vaccine dose? | Yes/No<br>1 \| Yes<br>0 \| No |
| In the last 7 days- what was the SEVERITY of your ACHING MUSCLES at their WORST? | dropdown<br>1 \| None<br>2 \| Mild |

| | |
|---|---|
| | 3 Moderate<br>4 Severe<br>5 Very severe |
| Section Header: *Section 3*<br>Did you have new or worsened joint pain since your vaccination? | Yes/No<br>1 Yes<br>0 No |
| If yes- In the last 7 days- how much did ACHING JOINTS (SUCH AS ELBOWS- KNEES- SHOULDERS) INTERFERE with your usual or daily activities? | dropdown<br>1 Not at all<br>2 A little bit<br>3 Somewhat<br>4 Quite a bit<br>5 Very much |
| Did you have new or worsened nausea since your vaccination? | Yes/No<br>1 Yes<br>0 No |
| In the last 7 days- how OFTEN did you have NAUSEA? | dropdown<br>1 Never<br>2 Rarely<br>3 Occasionally<br>4 Frequently<br>5 Almost constantly |
| In the last 7 days- what was the SEVERITY of your NAUSEA at its WORST? | dropdown<br>1 None<br>2 Mild<br>3 Moderate<br>4 Severe<br>5 Very severe |
| Did you have new or worsened vomiting since your vaccination? | Yes/No<br>1 Yes<br>0 No |
| In the last 7 days- what was the SEVERITY of your VOMITING at its WORST? | dropdown<br>1 None<br>2 Mild<br>3 Moderate<br>4 Severe<br>5 Very severe |

| | |
|---|---|
| Did you have new or worsened diarrhoea since your vaccination? | Yes/No- Required<br>1 Yes<br>0 No |
| In the last 7 days- how OFTEN did you have LOOSE OR WATERY STOOLS (DIARRHOEA)? | dropdown<br>1 Never<br>2 Rarely<br>3 Occasionally<br>4 Frequently<br>5 Almost constantly |
| How severe was your diarrhoea? | dropdown<br>1 Mild: 2 to 3 loose stools in 24 hours<br>2 Moderate: 4 to 5 loose stools in 24 hours<br>3 Severe: 6 or more loose stools in 24 hours<br>4 Very severe: I needed to attend the emergency department or I was admitted to hospital for diarrhoea |
| Section Header: *Section 4*<br>Have you had to delay or stop your anticancer treatment since your vaccination? | Yes/No<br>1 Yes<br>0 No |
| Have you attended the emergency department since your vaccination? | dropdown<br>0 No<br>1 Yes (Monash Health: Clayton- Dandenong or Casey Emergency Dept)<br>3 Yes (Westmead Hospital)<br>4 Yes (Sydney Children's Hospital)<br>2 Yes (other site- please specify where) |
| Which emergency department did you attend? | text (alpha_only) |
| Have you been admitted to hospital for any reason since your vaccination? | dropdown<br>1 Yes (Monash Health: Clayton (Monash Medical Centre)- Dandenong - Casey- Moorabbin- Kingston))<br>4 Yes (Westmead Hospital)<br>5 Yes (Sydney Children's Hospital)<br>2 Yes (other site- please specify where)<br>3 No |
| Which hospital were you admitted to? | text (alpha_only) |
| Have you started new treatment for a blood clot since your vaccination? | Yes/No- Required<br>1 Yes<br>0 No |

<table>
<tr><td>Have you needed to see your local doctor since vaccination?</td><td>dropdown<br><table><tr><td>1</td><td>Yes- routine check or prescription</td></tr><tr><td>2</td><td>Yes - unwell (please specify issue)</td></tr><tr><td>3</td><td>No</td></tr></table></td></tr>
<tr><td>Why did you see your local doctor?</td><td>text</td></tr>
<tr><td>Please describe any other side effect you had which has not been covered in the survey. Leave blank if nothing you need to add.</td><td>text</td></tr>
</table>

CAPITAL letters used for emphasis; capitalisation as per published PRO-CTCAE questionnaire item formatting.
