## Appendix 3 for "Vaccine beliefs, adverse effects, and quality of life in patients with cancer undergoing routine COVID-19 vaccination"

### Appendix 3. Results

**3a Proportion of patients responding to each survey**

Surveys were issued as per schedule in Figure 1.

| Activity completed | Adults<br>N=392 | Children (5-19yrs)*<br>N=107 |
|---|---|---|
| **Dose 1** | | |
| Received dose | 392 | 107 |
| Pre-dose QLQ-C30 | 379 | 4 |
| Pre-dose PedsQL | N/A | 45 |
| Pre-dose PedsQL proxy | N/A | 54 |
| Vaccine beliefs | 387 | 56 |
| Parent vaccine beliefs | N/A | 58 |
| Post-dose QLQ-C30 | 368 | 4 |
| Post-dose PedsQL | N/A | 39 |
| Post-dose PedsQL proxy | N/A | 55 |
| Patient-reported AEs | 371 | 54 |
| **Dose 2** | | |
| Received dose | 383 | 104 |
| Pre-dose QLQ-C30 | 369 | 5 |
| Pre-dose PedsQL | N/A | 45 |
| Pre-dose PedsQL proxy | N/A | 50 |
| Post-dose QLQ-C30 | 362 | 4 |
| Post-dose PedsQL | N/A | 32 |
| Post-dose PedsQL proxy | N/A | 46 |
| Patient-reported AEs | 367 | 28 |
| **Dose 3** | | |
| Received dose | 320 | 79 |
| Vaccine beliefs | 318 | 3 |
| Parent vaccine beliefs | N/A | 0 |
| Post-dose QLQ-C30 | 281 | 4 |
| Post-dose PedsQL | N/A | 19 |
| Post-dose PedsQL proxy | N/A | 29 |
| Patient-reported AEs | 288 | 27 |
| **Follow-up** | | |
| Follow-up QLQ-C30 | 312 | 3 |
| Follow-up PedsQL | N/A | 38 |
| Follow-up PedsQL proxy | N/A | 0 |
| Medical record review | 388 | 104 |

N = number; N/A = not applicable

* 18 and 19 year-olds received either QLQ-C30 or PedsQL questionnaires depending if enrolled at an adult or children's cancer centre.

**3b Vaccine related beliefs- Oxford Complacency and Confidence Scale- Adults**

| | Dose 1 (N = 387) | Dose 3 (N = 318) |
|---|---|---|
| **Importance** | | |
| N | 357 | 269 |
| Mean (SD) | 17.0 (10.6) | 18.4 (12.6) |
| Median (IQR) | 15.0 (10.0 - 25.0) | 18.8 (10.0 - 25.0) |
| Unknown (%) | 30 (8) | 49 (15) |
| **Efficacy** | | |
| N | 336 | 267 |
| Mean (SD) | 37.2 (13.7) | 36.5 (14.3) |
| Median (IQR) | 37.5 (33.3 - 50.0) | 37.5 (25.0 - 50.0) |
| Unknown (%) | 51 (13) | 51 (16) |
| **Speed of development** | | |
| N | 296 | 0 |
| Mean (SD) | 24.0 (16.0) | N/A |
| Median (IQR) | 25.0 (12.5 - 33.3) | N/A |
| Unknown (%) | 91 (24) | N/A |
| **Side effects** | | |
| N | 353 | 277 |
| Mean (SD) | 21.0 (14.6) | 19.4 (16.1) |
| Median (IQR) | 16.7 (8.3 - 25.0) | 16.7 (8.3 - 25.0) |
| Unknown (%) | 34 (9) | 41 (13) |
| **Total score** | | |
| N | 255 | 0 |
| Mean (SD) | 24.0 (9.3) | N/A |
| Median (IQR) | 23.8 (18.0 - 28.9) | N/A |
| Unknown (%) | 132 (34) | N/A |
| **Total score*** | | |
| N | 309 | 234 |
| Mean (SD) | 24.7 (9.5) | 23.9 (10.3) |
| Median (IQR) | 23.9 (18.9 - 30.0) | 22.8 (17.2 - 29.2) |
| Unknown (%) | 78 (20) | 84 (26) |

N = number N/A = not applicable at this timepoint; SD= Standard Deviation; IQR= Interquartile Range
*excluded question regarding speed of vaccine development

**3c Vaccine related beliefs- Oxford Complacency and Confidence Scale- Children**

| | Patient-reported (N = 56) | Parent-reported (N = 58) |
|---|---|---|
| **Importance** | | |
| N | 53 | 56 |
| Mean (SD) | 16.8 (9.4) | 16.3 (10.2) |
| Median (IQR) | 15.0 (10.0 - 25.0) | 15.0 (10.0 - 25.0) |
| Unknown (%) | 3 (5) | 2 (3) |
| **Efficacy** | | |
| N | 51 | 57 |
| Mean (SD) | 35.3 (14.4) | 31.9 (12.5) |
| Median (IQR) | 37.5 (25.0 - 50.0) | 33.3 (25.0 - 41.7) |
| Unknown (%) | 5 (9) | 1 (2) |
| **Speed of development** | | |
| N | 43 | 47 |
| Mean (SD) | 26.2 (15.9) | 31.0 (16.7) |
| Median (IQR) | 25.0 (16.7 - 33.3) | 33.3 (16.7 - 50.0) |
| Unknown (%) | 13 (23) | 11 (19) |
| **Side effects** | | |
| N | 50 | 56 |
| Mean (SD) | 20.9 (13.0) | 20.8 (11.6) |
| Median (IQR) | 16.7 (12.5 - 25.0) | 16.7 (16.7 - 25.0) |
| Unknown (%) | 6 (11) | 2 (3) |
| **Total score** | | |
| N | 40 | 47 |
| Mean (SD) | 24.0 (8.0) | 25.0 (8.7) |
| Median (IQR) | 23.5 (18.2 - 29.2) | 24.6 (20.8 - 28.9) |
| Unknown (%) | 16 (29) | 11 (19) |
| **Total score*** | | |
| N | 49 | 56 |
| Mean (SD) | 24.1 (8.2) | 22.9 (8.6) |
| Median (IQR) | 23.3 (18.9 - 27.8) | 21.8 (16.9 - 26.8) |
| Unknown (%) | 7 (12) | 2 (3) |

N = number N/A = not applicable at this timepoint; SD = Standard Deviation; IQR= Interquartile Range

*excluded question regarding speed of vaccine development

**3d Grade and attribution of investigator-reported Serious Adverse Events**

Serious adverse events (SAEs) recorded from medical record reviews were graded according to Common Terminology Criteria for Adverse Events (CTCAE) Version 5.0.

| | Adults n (%- 95% CI) | | Children n (%- 95% CI) | |
|---|---|---|---|---|
| | **Dose 1-2*** | **Dose 3**** | **Dose 1-2*** | **Dose 3**** |
| **Any SAE** | | | | |
| No | 337 (88; 85-91) | 295 (97; 95-99) | 71 (70; 60-78) | 51 (81; 69-90) |
| Yes | 45 (12; 9-15) | 8 (3; 1-5) | 31 (30; 22-40) | 12 (19; 10-31) |
| **Number of SAE** | | | | |
| 0 | 337 (88; 85-91) | 295 (97; 95-99) | 71 (70; 60-78) | 51 (81; 69-90) |
| 1 | 31 (8; 6-11) | 7 (2; 1-5) | 20 (20; 12-29) | 10 (16; 8-27) |
| 2 | 11 (3; 1-5) | 1 (0; 0-2) | 8 (8; 3-15) | 2 (3; 0-11) |
| 3 | 2 (1; 0-2) | 0 | 2 (2; 0-7) | 0 |
| 4+ | 1 (0; 0-1) | 0 | 1 (1; 0-5) | 0 |
| **Worst grade** | | | | |
| 1 | 3 (1; 0-2) | 0 (0; 0-1) | 4 (4; 1-10) | 6 (10; 4-20) |
| 2 | 6 (2; 1-3) | 0 (0; 0-1) | 3 (3; 1-8) | 4 (6; 2-15) |
| 3 | 32 (8; 6-12) | 8 (3; 1-5) | 21 (21; 13-30) | 2 (3; 0-11) |
| 4 | 2 (1; 0-2) | 0 (0; 0-1) | 2 (2; 0-7) | 0 |
| 5 | 2 (1; 0-2) | 0 | 0 | 0 |
| **Attributable to vaccine** | | | | |
| Unlikely | 41 (11; 8-14) | 8 (3; 1-5) | 17 (17; 10-25) | 6 (10; 4-20) |
| Possible | 4 (1; 0-3) | 0 (0; 0-1) | 14 (14; 8-22) | 3 (5; 1-13) |
| Attributed | 0 | 0 | 0 | 3 (5; 1-13) |

Time period relates to *dose 1 until one month post-dose 2 and **dose 3 until one month after.
SAE = Serious Adverse Event; N = number; CI = confidence interval

**3e Investigator-determined attribution of Serious Adverse Events**

| | Adults | Children |
|---|---|---|
| **Definitely or very likely related** | | |
| Fever | 0 | 2 |
| Headache | 0 | 1 |
| **Possibly related** | | |
| Fever | 1 | 20 |
| Mucositis | 0 | 2 |
| Rash | 1 | 0 |
| Upper respiratory symptoms | 1 | 0 |
| Pulmonary embolism | 1* | 0 |
| Stroke | 1* | 0 |

* Same participant, who suffered pulmonary embolism 5 days post initial BNT162b2 vaccine then an ischemic stroke 7 days later, in the context of untreated malignancy and intercurrent pneumonia.

*Note: One adult with comorbid ischaemic heart disease had a fatal myocardial infarction six weeks after the second BNT162b2 dose, outside the window for SAE reporting.*

**3f Category of Participant reported adverse events**

Participant reported adverse events (AE) were recorded according to selected questions from PROCTCAE (Appendix 1c).

| | | Adults | | Children | |
|---|---|---|---|---|---|
| | Dose | n* / N | % (95% CI) | n / N | % (95% CI) |
| **Local side effect (at injection site)** | | | | | |
| | Dose 1 | 240 / 369 | 65 (60-70) | 46 / 54 | 85 (73-93) |
| | 2 | 245 / 365 | 67 (62-72) | 19 / 28 | 68 (48-84) |
| | 3 | 196 / 286 | 69 (63- 74) | 21 / 27 | 78 (58-91) |
| **Systemic side effect** | | | | | |
| | Dose 1 | 178 / 353 | 50 (45-56) | 26 / 50 | 52 (37-66) |
| | 2 | 217 / 351 | 62 (57-67) | 16 / 27 | 59 (39-78) |
| | 3 | 159 / 273 | 58 (52-64) | 14 / 26 | 54 (33-73) |
| **Other side effect or adverse event** | | | | | |
| | Dose 1 | 108 / 371 | 29 (25-34) | 18 / 54 | 33 (21-47) |
| | 2 | 117 / 366 | 32 (27-37) | 8 / 28 | 29 (13-49) |
| | 3 | 102 / 288 | 35 (30-41) | 10 / 26 | 38 (20-59) |
| **Healthcare utilisation** | | | | | |
| | Dose 1 | 29 / 315 | 9 (6-13) | 13 / 53 | 25 (14-38) |
| | 2 | 35 / 347 | 10 (7-14) | 2 / 26 | 8 (1- 25) |
| | 3 | 17 / 274 | 6 (4-10) | 8 / 26 | 31 (14-52) |
| **Delay or interruption to cancer treatment** | | | | | |
| | Dose 1 | 6 / 370 | 2 (1-3) | 4 / 54 | 7 (2-18) |
| | 2 | 14 / 365 | 4 (2-6) | 0 / 27 | 0 (0-13) |
| | 3 | 6 / 288 | 2 (1-4) | 1 / 26 | 4 (0-20) |

*n= number reporting an adverse effect; N=total number of respondents at timepoint; CI = Confidence Interval
**unscheduled visit to doctor or hospital

**3g Specific Participant reported adverse events**

| | | Adults | | Children | |
|---|---|---|---|---|---|
| | **Dose** | **n/N** | **% (95% CI)** | **n/N** | **% (95% CI)** |
| **Pain at injection site** | | | | | |
| | Dose 1 | 224 / 371 | 60 (55- 65) | 45 / 54 | 83 (71- 92) |
| | 2 | 229 / 367 | 62 (57- 67) | 17 / 28 | 61 (41- 78) |
| | 3 | 180 / 288 | 62 (57- 68) | 20 / 27 | 74 (54- 89) |
| **Medication for side effects** | | | | | |
| | Dose 1 | 105 / 371 | 28 (24- 33) | 17 / 54 | 31 (20- 46) |
| | 2 | 116 / 367 | 32 (27- 37) | 8 / 28 | 29 (13- 49) |
| | 3 | 100 / 288 | 35 (29- 41) | 10 / 27 | 37 (19- 58) |
| **Fatigue** | | | | | |
| | Dose 1 | 75 / 371 | 20 (16- 25) | 11 / 54 | 20 (11- 34) |
| | 2 | 114 / 367 | 31 (26- 36) | 12 / 28 | 43 (24- 63) |
| | 3 | 83 / 288 | 29 (24- 34) | 7 / 26 | 27 (12- 48) |
| **Muscle pain** | | | | | |
| | Dose 1 | 56 / 367 | 15 (12- 19) | 9 / 53 | 17 (8- 30) |
| | 2 | 87 / 363 | 24 (20- 29) | 5 / 28 | 18 (6- 37) |
| | 3 | 72 / 283 | 25 (20- 31) | 4 / 26 | 15 (4- 35) |
| **Headache** | | | | | |
| | Dose 1 | 66 / 371 | 18 (14- 22) | 9 / 54 | 17 (8- 29) |
| | 2 | 80 / 367 | 22 (18- 26) | 6 / 28 | 21 (8- 41) |
| | 3 | 56 / 288 | 19 (15- 24) | 4 / 26 | 15 (4- 35) |
| **Redness or swelling at injection site** | | | | | |
| | Dose 1 | 48 / 367 | 13 (10- 17) | 9 / 54 | 17 (8- 20) |
| | 2 | 63 / 363 | 17 (14- 22) | 1 / 28 | 4 (0- 18) |
| | 3 | 62 / 283 | 22 (17- 27) | 3 / 27 | 11 (2- 29) |

| | | Adults | | Children | |
|---|---|---|---|---|---|
| | Dose | n/N | % (95% CI) | n/N | % (95% CI) |
| **Shivering or shaking chills** | | | | | |
| | Dose 1 | 44 / 362 | 12 (9- 16) | 8 / 52 | 15 (7- 28) |
| | 2 | 57 / 354 | 16 (12- 20) | 7 / 27 | 26 (11- 46) |
| | 3 | 53 / 277 | 19 (15- 24) | 5 / 26 | 19 (7- 39) |
| **Joint pain** | | | | | |
| | Dose 1 | 40 / 371 | 11 (8- 14) | 5 / 54 | 9 (3- 20) |
| | 2 | 63 / 367 | 17 (13- 21) | 1 / 28 | 4 (0- 18) |
| | 3 | 62 / 288 | 22 (17- 27) | 0 / 26 | 0 (0- 13) |
| **Itch at injection site** | | | | | |
| | Dose 1 | 44 / 371 | 12 (9- 16) | 10 / 54 | 19 (9- 31) |
| | 2 | 57 / 367 | 16 (12- 20) | 4 / 28 | 14 (4- 33) |
| | 3 | 47 / 288 | 16 (12- 21) | 6 / 27 | 22 (9- 42) |
| **Fever** | | | | | |
| | Dose 1 | 27 / 351 | 8 (5- 11) | 6 / 50 | 12 (5- 24) |
| | 2 | 54 / 351 | 15 (12- 20) | 3 / 25 | 12 (3- 31) |
| | 3 | 56 / 277 | 20 (16- 25) | 4 / 26 | 15 (4- 35) |
| **Nausea** | | | | | |
| | Dose 1 | 27 / 369 | 7 (5- 10) | 4 / 54 | 7 (2- 18) |
| | 2 | 39 / 367 | 11 (8- 14) | 5 / 28 | 18 (6- 37) |
| | 3 | 21 / 285 | 7 (5- 11) | 4 / 26 | 15 (4- 35) |
| **Diarrhoea** | | | | | |
| | Dose 1 | 17 / 371 | 5 (3- 7) | 5 / 54 | 9 (3- 20) |
| | 2 | 26 / 367 | 7 (5- 10) | 0 / 28 | 0 (0- 12) |
| | 3 | 21 / 288 | 7 (5- 11) | 2 / 26 | 8 (1- 25) |

| | | Adults | | Children | |
|---|---|---|---|---|---|
| | **Dose** | **n/N** | **% (95% CI)** | **n/N** | **% (95% CI)** |
| **Hospital admission** | | | | | |
| | Dose 1 | 20 / 339 | 6 (4- 9) | 9 / 52 | 17 (8- 30) |
| | 2 | 16 / 347 | 5 (3- 7) | 2 / 26 | 8 (1- 25) |
| | 3 | 8 / 273 | 3 (1- 6) | 5 / 26 | 19 (7- 39) |
| **Other systemic side effect** | | | | | |
| | Dose 1 | 13 / 371 | 4 (2- 6) | 13 / 371 | 4 (2- 6) |
| | 2 | 19 / 367 | 5 (3- 8) | 19 / 367 | 5 (3- 8) |
| | 3 | 15 / 288 | 5 (3- 8) | 15 / 288 | 5 (3- 8) |
| **Emergency department attendance** | | | | | |
| | Dose 1 | 10 / 330 | 3 (1- 6) | 7 / 54 | 13 (5- 25) |
| | 2 | 15 / 364 | 4 (2- 7) | 1 / 27 | 4 (0- 19) |
| | 3 | 3 / 286 | 1 (0- 3) | 6 / 26 | 23 (9- 44) |
| **Local doctor visit** | | | | | |
| | Dose 1 | 10 / 365 | 3 (1- 5) | 2 / 53 | 4 (0- 13) |
| | 2 | 15 / 363 | 4 (2- 7) | 1 / 27 | 4 (0- 19) |
| | 3 | 7 / 285 | 2 (1- 5) | 0 / 26 | 0 (0- 13) |
| **Delays or interruption to anticancer treatment** | | | | | |
| | Dose 1 | 6 / 370 | 2 (1- 3) | 4 / 54 | 7 (2- 18) |
| | 2 | 14 / 365 | 4 (2- 6) | 0 / 27 | 0 (0- 13) |
| | 3 | 6 / 288 | 2 (1- 4) | 1 / 26 | 4 (0- 20) |
| **Vomiting** | | | | | |
| | Dose 1 | 3 / 369 | 1 (0- 2) | 3 / 54 | 6 (1- 15) |
| | 2 | 11 / 366 | 3 (2- 5) | 2 / 28 | 7 (1- 24) |
| | 3 | 5 / 286 | 2 (1- 4) | 2 / 26 | 8 (1- 25) |

| | | Adults | | Children | |
|---|---|---|---|---|---|
| | Dose | n/N | % (95% CI) | n/N | % (95% CI) |
| **Other local side effect** | | | | | |
| | Dose 1 | 1 / 371 | 0 (0- 1) | 1 / 54 | 2 (0- 10) |
| | 2 | 3 / 367 | 1 (0- 2) | 0 / 28 | 0 (0- 12) |
| | 3 | 2 / 288 | 1 (0- 2) | 1 / 27 | 4 (0- 19) |
| **Treatment for blood clot** | | | | | |
| | Dose 1 | 3 / 371 | 1 (0- 2) | 1 / 54 | 2 (0- 10) |
| | 2 | 1 / 366 | 0 (0- 2) | 0 / 27 | 0 (0- 13) |
| | 3 | 2 / 288 | 1 (0- 2) | 0 / 26 | 0 (0- 13) |
| **Other side effect** | | | | | |
| | Dose 1 | 0 / 371 | 0 (0- 1) | 0 / 54 | 0 (0- 7) |
| | 2 | 0 / 367 | 0 (0- 1) | 0 / 28 | 0 (0- 12) |
| | 3 | 0 / 288 | 0 (0- 1) | 0 / 27 | 0 (0- 13) |

*n = number reporting an adverse effect; N = total number of respondents at timepoint; CI = Confidence Interval ED = Emergency Department

**unscheduled visit to doctor or hospital

**3h Association of worst patient reported adverse event with receipt of 3rd vaccine dose**

| Worst side effect* | Received 3rd dose n / N (%) | OR (95% CI) |
|---|---|---|
| None | 37 / 48 (77) | — |
| Mild | 124 / 148 (84) | 1.54 (0.7- 3.4) |
| Moderate | 98 / 108 (91) | 2.91 (1.1- 7.6) |
| Severe | 48 / 58 (83) | 1.43 (0.6- 3.8) |
| Serious | 11 / 18 (61) | 0.47 (0.2- 1.5) |

Multiple imputation was conducted to account for missing data.
The severity of participant-reported AE after the first two vaccinations was not significantly associated with receipt of a third dose [p=0.48]
*Severity self-reported by 5-point Likert scale (Appendix 1b).
n = number reporting an adverse effect; N = total number of respondents at timepoint; OR = Odds Ratio; CI = Confidence Interval

**3i Investigator- reported adverse events by vaccine type**

Incidence of investigator-reported adverse events observed between dose 1 and one month post-dose 2; and between dose 3 and 1 month post dose 3, for patients over 60 years, comparing those who received an mRNA vaccine (BNT162b2 or mRNA-1273) and those who received the viral vector (ChadOx1-S) vaccine. 1 patient received a different vaccine for dose 1 and 2 and was excluded.

| Dose | Events / Patients n♦ / N (%) mRNA | Viral vector | OR+ (95% CI) | p-value | RR+ + (95% CI) | p-value |
|---|---|---|---|---|---|---|
| **Thrombotic event** | | | | | | |
| Dose 1 - 2* | 2 / 134 (1) | 0 / 37 (0) | NR | 0.32 | NR | 0.22 |
| 3** | 0 / 141 (0) | 0 / 2 (0) | NR | — | NR | — |
| **Allergic reaction** | | | | | | |
| Dose 1 - 2 | 0 / 134 (0) | 0 / 37 (0) | NR | — | NR | — |
| 3 | 0 / 141 (0) | 0 / 2 (0) | NR | — | NR | — |
| **Decline in performance status*** ** | | | | | | |
| Dose 1 - 2 | 7 / 132 (5) | 4 / 37 (11) | 2.16 (0.5 - 7.6) | 0.26 | 1.21 (0.3 - 4.0) | 0.76 |
| 3 | 6 / 134 (4) | 0 / 2 (0) | NR | 0.67 | NR | 0.67 |
| **Lymphadenopathy** | | | | | | |
| Dose 1 - 2 | 8 / 134 (6) | 5 / 37 (14) | 2.46 (0.7 - 7.9) | 0.15 | 1.35 (0.4 - 4.0) | 0.60 |
| 3 | 3 / 141 (2) | 0 / 2 (0) | NR | 0.77 | NR | 0.77 |
| **Treatment delay** | | | | | | |
| Dose 1 - 2 | 8 / 134 (6) | 4 / 37 (11) | 1.91 (0.5 - 6.5) | 0.33 | 1.08 (0.3 - 3.4) | 0.90 |
| 3 | 7 / 141 (5) | 0 / 2 (0) | NR | 0.65 | NR | 0.66 |
| **Treatment modification** | | | | | | |
| Dose 1 - 2 | 21 / 134 (16) | 5 / 37 (14) | 0.84 (0.3 - 2.3) | 0.74 | 0.51 (0.2 - 1.3) | 0.15 |
| 3 | 15 / 141 (11) | 0 / 2 (0) | NR | 0.50 | NR | 0.52 |
| **Treatment delay and/or modification** | | | | | | |
| Dose 1 - 2 | 23 / 134 (17) | 7 / 37 (19) | 1.13 (0.4 - 2.8) | 0.81 | 0.66 (0.3 - 1.5) | 0.31 |
| 3 | 20 / 141 (14) | 0 / 2 (0) | NR | 0.44 | NR | 0.45 |

* between dose 1 and one month post-dose 2; ** between dose 3 and one month afterwards; *** Eastern Cooperative Group performance status
+ OR=odds ratio; estimated from a logistic regression model for each adverse event at each time point; + + RR = relative rate: as the period between doses 1 and 2 differed by vaccine type, a log-link Poisson GLM including the log of the actual time period as an offset was fitted to estimate the relative rate.
NR = not recordable as no events observed in one or both vaccine groups.
Allergic reactions were recorded separately for each dose; we have combined doses 1 & 2 here for consistency
♦n = number reporting an adverse effect; N = total number of respondents at timepoint CI = Confidence Interval

##### 3j Serious Adverse Events by vaccine type

The number of SAEs at during each reporting period were compared between vaccine types, relative rate was adjusted according to exposure time.

| | **SAEs / Patients ((SAE rate per 30 person days)#)** | | **Unadjusted** | | **Adjusted*** | |
|---|---|---|---|---|---|---|
| **Dose** | **mRNA** (BNT162b2) | **Viral vector** (ChAdOx1-S) | **RR (95% CI)** | **p-value** | **RR (95% CI)** | **p-value** |
| 1 and 2 | 12 / 135 (0.09) | 10 / 37 (0.27) | 3.04 (1.28, 7.05) | 0.013 | 1.81 (0.76, 4.20) | 0.17 |
| 3 | 4 / 140 (0.03) | 0 / 2 (0.00) | — | 0.74 | — | 0.74 |

#Fitted rate of SAEs per 30 person-days under unadjusted log-link Poisson generalised linear model (GLM),

*exposure time was adjusted for by adjusted log-link Poisson GLM, to account for differences in dosing schedule and subsequent time at risk during each reporting period. Median exposure time from dose 1 to 1 month post dose 2 was 51 days for mRNA recipients and 80 days for viral vector recipients.

Adj. Adjusted (but I think just simplify the table heading to adjusted)

Exp. Exp. exposure (but as above, covered in the footer)

CI Confidence Interval

RR Relative Rate

.

**3k Participant-reported adverse effects by vaccine type**

Participant-reported side effects were measured by selected PROCTCAE items (Appendix 1c) in patients over 60 years to compare mRNA and viral vector vaccine types.

| Events / Respondents (%) by vaccine type | | |
|---|---|---|
| | mRNA | Viral vector |
| **Pain at injection site** | | |
| Dose 1 | 74 / 135 (55) | 15 / 39 (38) |
| 2 | 71 / 134 (53) | 8 / 35 (23) |
| 3 | 71 / 133 (53) | 1 / 2 (50) |
| **Redness or swelling at injection site** | | |
| Dose 1 | 14 / 132 (11) | 5 / 39 (13) |
| 2 | 16 / 133 (12) | 3 / 34 (9) |
| 3 | 25 / 130 (19) | 2 / 2 (100) |
| **Itch at injection site** | | |
| Dose 1 | 17 / 135 (13) | 5 / 39 (13) |
| 2 | 16 / 134 (12) | 3 / 35 (9) |
| 3 | 22 / 133 (17) | 0 / 2 (0) |
| **Medication for side effects** | | |
| Dose 1 | 31 / 135 (23) | 8 / 39 (21) |
| 2 | 32 / 134 (24) | 4 / 35 (11) |
| 3 | 36 / 133 (27) | 1 / 2 (50) |
| **Fatigue** | | |
| Dose 1 | 24 / 135 (18) | 4 / 39 (10) |
| 2 | 33 / 134 (25) | 5 / 35 (14) |
| 3 | 35 / 133 (26) | 0 / 2 (0) |
| **Muscle pain** | | |
| Dose 1 | 21 / 134 (16) | 5 / 39 (13) |
| 2 | 24 / 133 (18) | 6 / 33 (18) |
| 3 | 28 / 129 (22) | 0 / 2 (0) |
| **Joint pain** | | |
| Dose 1 | 14 / 135 (10) | 6 / 39 (15) |
| 2 | 20 / 134 (15) | 6 / 35 (17) |
| 3 | 30 / 133 (23) | 2 / 2 (100) |

| Events / Respondents (%) by vaccine type | | | |
|---|---|---|---|
| | | mRNA | Viral vector |
| **Shivering or shaking chills** | | | |
| | Dose 1 | 14 / 131 (11) | 7 / 38 (18) |
| | 2 | 15 / 130 (12) | 6 / 33 (18) |
| | 3 | 24 / 125 (19) | 1 / 2 (50) |
| **Headache** | | | |
| | Dose 1 | 20 / 135 (15) | 5 / 39 (13) |
| | 2 | 22 / 134 (16) | 3 / 35 (9) |
| | 3 | 17 / 133 (13) | 2 / 2 (100) |
| **Fever** | | | |
| | Dose 1 | 10 / 126 (8) | 1 / 38 (3) |
| | 2 | 12 / 126 (10) | 1 / 32 (3) |
| | 3 | 20 / 127 (16) | 0 / 2 (0) |
| **Nausea** | | | |
| | Dose 1 | 11 / 134 (8) | 3 / 39 (8) |
| | 2 | 11 / 134 (8) | 3 / 35 (9) |
| | 3 | 11 / 132 (8) | 0 / 2 (0) |
| **Diarrhoea** | | | |
| | Dose 1 | 9 / 135 (7) | 1 / 39 (3) |
| | 2 | 9 / 134 (7) | 2 / 35 (6) |
| | 3 | 8 / 133 (6) | 0 / 2 (0) |
| **Hospital admission** | | | |
| | Dose 1 | 7 / 125 (6) | 0 / 33 (0) |
| | 2 | 4 / 125 (3) | 2 / 31 (6) |
| | 3 | 4 / 123 (3) | 0 / 2 (0) |
| **Local doctor visit** | | | |
| | Dose 1 | 6 / 132 (5) | 0 / 39 (0) |
| | 2 | 6 / 132 (5) | 2 / 34 (6) |
| | 3 | 4 / 131 (3) | 0 / 2 (0) |
| **Other systemic side effect** | | | |
| | Dose 1 | 5 / 135 (4) | 1 / 39 (3) |
| | 2 | 4 / 134 (3) | 1 / 35 (3) |
| | 3 | 2 / 133 (2) | 1 / 2 (50) |
| **Delays in interruption to anticancer treatment** | | | |
| | Dose 1 | 2 / 134 (1) | 0 / 39 (0) |
| | 2 | 5 / 133 (4) | 2 / 35 (6) |
| | 3 | 4 / 133 (3) | 0 / 2 (0) |

| | Events / Respondents (%) by vaccine type | |
|---|---|---|
| | **mRNA** | **Viral vector** |
| **Vomiting** | | |
| Dose 1 | 1 / 135 (1) | 0 / 37 (0) |
| 2 | 5 / 134 (4) | 2 / 35 (6) |
| 3 | 3 / 133 (2) | 0 / 2 (0) |
| **Emergency department attendance** | | |
| Dose 1 | 2 / 127 (2) | 0 / 29 (0) |
| 2 | 5 / 132 (4) | 1 / 34 (3) |
| 3 | 1 / 132 (1) | 0 / 2 (0) |
| **Treatment for blood clot** | | |
| Dose 1 | 0 / 135 (0) | 2 / 39 (5) |
| 2 | 0 / 133 (0) | 1 / 35 (3) |
| 3 | 2 / 133 (2) | 0 / 2 (0) |
| **Other local side effect** | | |
| Dose 1 | 0 / 135 (0) | 0 / 39 (0) |
| 2 | 0 / 134 (0) | 2 / 35 (6) |
| 3 | 0 / 133 (0) | 0 / 2 (0) |

**3l Impact of sequential vaccine doses on health-related quality of life in adults**

Quality of life measured using the European Organization for Research and Treatment of Cancer Quality of Life Core Questionnaire 30 item instrument (EORTC QLQC-30). Multiple imputations were completed to account for missing data.

| | | Observed | | | | Imputed | | | | | |
|---|---|---|---|---|---|---|---|---|---|---|---|
| | | Pre | | Post | | Pre | | Post | | Difference | |
| | **Dose** | **N** | **Mean (SD)** | **N** | **Mean (SD)** | **N** | **Mean (SD)** | **N** | **Mean (SD)** | **Est (95% CI)** | **p-value** |
| **Primary outcomes** | | | | | | | | | | | |
| Global health status / QoL | 1 | 377 | 68.8 (20.5) | 366 | 67.3 (22.2) | 386 | 68.8 (20.6) | 386 | 67.4 (22.1) | -1.4 (-3.4 - 0.6) | 0.16 |
| | 2 | 368 | 65.9 (22.6) | 361 | 66.9 (21.5) | 378 | 66.2 (22.6) | 378 | 66.9 (21.6) | 0.7 (-1.2 - 2.6) | 0.48 |
| Summary score | 1 | 369 | 82.4 (14.3) | 360 | 83.0 (14.5) | 384 | 82.4 (14.4) | 384 | 82.9 (14.7) | 0.5 (-0.5 - 1.6) | 0.30 |
| | 2 | 362 | 82.2 (15.4) | 359 | 81.8 (16.7) | 377 | 82.6 (15.3) | 377 | 81.8 (16.6) | -0.8 (-1.9 - 0.3) | 0.17 |
| **Functional scales** | | | | | | | | | | | |
| Physical functioning | 1 | 379 | 81.7 (19.7) | 368 | 82.2 (18.9) | 387 | 81.6 (19.8) | 387 | 82.2 (18.9) | 0.7 (-0.7 - 2.1) | 0.35 |
| | 2 | 369 | 81.1 (20.9) | 362 | 81.9 (21.0) | 378 | 81.3 (20.8) | 378 | 82.0 (21.0) | 0.7 (-0.9 - 2.2) | 0.40 |
| Role functioning | 1 | 379 | 76.5 (27.3) | 367 | 77.8 (26.7) | 387 | 76.4 (27.3) | 387 | 77.7 (26.8) | 1.3 (-1.2 - 3.8) | 0.29 |
| | 2 | 369 | 77.7 (27.2) | 362 | 76.6 (29.1) | 378 | 78.0 (27.1) | 378 | 76.7 (28.9) | -1.3 (-3.7 - 1.1) | 0.28 |
| Emotional functioning | 1 | 377 | 80.4 (19.4) | 367 | 81.2 (20.2) | 386 | 80.2 (19.6) | 386 | 81.3 (20.2) | 1.1 (-0.5 - 2.8) | 0.19 |
| | 2 | 368 | 80.2 (20.1) | 361 | 79.8 (22.8) | 378 | 80.4 (20.1) | 378 | 79.9 (22.7) | -0.5 (-2.1 - 1.1) | 0.56 |
| Cognitive functioning | 1 | 377 | 85.0 (19.6) | 367 | 84.8 (20.4) | 386 | 84.8 (19.8) | 386 | 84.9 (20.2) | 0.0 (-1.8 - 1.8) | 0.97 |
| | 2 | 369 | 83.5 (20.7) | 361 | 82.8 (22.1) | 378 | 83.7 (20.6) | 378 | 82.8 (22.1) | -0.9 (-2.8 - 1.0) | 0.34 |
| Social functioning | 1 | 376 | 80.0 (26.0) | 367 | 81.4 (26.5) | 386 | 79.9 (26.0) | 386 | 81.4 (26.5) | 1.5 (-1.1 - 4.0) | 0.25 |
| | 2 | 368 | 80.6 (27.4) | 361 | 79.6 (28.7) | 378 | 80.9 (27.2) | 378 | 79.6 (28.6) | -1.3 (-3.7 - 1.0) | 0.26 |

| Symptom scales /items | | | | | | | | | | | |
|---|---|---|---|---|---|---|---|---|---|---|---|
| Fatigue | 1 | 377 | 32.2 (24.5) | 367 | 31.4 (24.7) | 386 | 32.4 (24.5) | 386 | 31.4 (24.6) | -1.0 (-3.1 - 1.2) | 0.39 |
| | 2 | 369 | 33.3 (24.5) | 361 | 33.7 (26.7) | 378 | 33.0 (24.4) | 378 | 33.7 (26.7) | 0.6 (-1.4 - 2.7) | 0.54 |
| Nausea & vomiting | 1 | 377 | 6.1 (12.9) | 366 | 5.9 (12.7) | 386 | 6.3 (13.3) | 386 | 6.0 (12.8) | -0.4 (-1.7 - 1.0) | 0.58 |
| | 2 | 369 | 6.5 (13.7) | 361 | 6.7 (13.2) | 378 | 6.5 (13.7) | 378 | 6.8 (13.2) | 0.3 (-1.0 - 1.5) | 0.66 |
| Pain | 1 | 379 | 20.2 (23.5) | 367 | 22.8 (25.6) | 387 | 20.2 (23.6) | 387 | 22.8 (25.5) | 2.6 (0.4 - 4.7) | 0.020 |
| | 2 | 369 | 21.7 (26.8) | 362 | 24.2 (27.5) | 378 | 21.5 (26.7) | 378 | 24.2 (27.5) | 2.7 (0.4 - 5.0) | 0.021 |
| Dyspnoea | 1 | 378 | 13.9 (21.3) | 366 | 12.4 (20.2) | 387 | 13.9 (21.2) | 387 | 12.3 (20.2) | -1.6 (-3.6 - 0.4) | 0.11 |
| | 2 | 369 | 15.3 (22.6) | 362 | 15.1 (22.3) | 378 | 15.0 (22.5) | 378 | 15.2 (22.3) | 0.1 (-1.8 - 2.1) | 0.89 |
| Insomnia | 1 | 376 | 25.0 (27.0) | 366 | 23.6 (28.1) | 386 | 25.3 (27.3) | 386 | 23.3 (27.9) | -2.0 (-4.4 - 0.4) | 0.096 |
| | 2 | 369 | 23.6 (26.4) | 360 | 23.8 (27.5) | 378 | 23.4 (26.3) | 378 | 23.7 (27.6) | 0.3 (-2.2 - 2.8) | 0.80 |
| Appetite loss | 1 | 375 | 13.5 (25.1) | 364 | 13.1 (22.5) | 385 | 13.4 (25.0) | 385 | 13.1 (22.4) | -0.4 (-2.6 - 1.9) | 0.75 |
| | 2 | 367 | 13.2 (23.6) | 360 | 14.2 (24.2) | 378 | 12.9 (23.4) | 378 | 14.1 (24.1) | 1.2 (-0.8 - 3.2) | 0.25 |
| Constipation | 1 | 375 | 12.1 (23.0) | 365 | 13.4 (23.8) | 386 | 12.2 (23.2) | 386 | 13.1 (23.6) | 1.0 (-1.3 - 3.2) | 0.39 |
| | 2 | 366 | 11.2 (21.2) | 361 | 12.0 (22.0) | 378 | 11.1 (21.1) | 378 | 12.0 (22.1) | 0.9 (-1.2 - 3.1) | 0.40 |
| Diarrhoea | 1 | 377 | 8.1 (18.2) | 366 | 7.1 (17.6) | 386 | 8.2 (18.1) | 386 | 7.0 (17.5) | -1.1 (-3.0 - 0.7) | 0.23 |
| | 2 | 366 | 9.4 (20.2) | 361 | 7.9 (18.9) | 378 | 9.2 (20.0) | 378 | 8.3 (19.6) | -0.9 (-2.9 - 1.0) | 0.35 |
| Financial difficulties | 1 | 375 | 14.2 (26.1) | 367 | 13.4 (26.3) | 386 | 14.4 (26.3) | 386 | 13.5 (26.6) | -0.8 (-2.9 - 1.2) | 0.41 |
| | 2 | 368 | 13.5 (25.8) | 361 | 12.7 (25.8) | 378 | 13.3 (25.6) | 378 | 12.8 (25.9) | -0.5 (-2.3 - 1.4) | 0.61 |

N = number; SD = Standard Deviation; QoL = Quality of Life; Est = Estimated; CI= Confidence Interval

**3m Impact of sequential doses on health-related quality of life in children**

i. **Child reported questionnaires**

| | | Observed | | | | Imputed | | | | | |
|---|---|---|---|---|---|---|---|---|---|---|---|
| | | Pre | | Post | | Pre | | Post | | Difference | |
| | **Dose** | **N** | **Mean (SD)** | **N** | **Mean (SD)** | **N** | **Mean (SD)** | **N** | **Mean (SD)** | **Est (95% CI)** | **p-value** |
| **Primary outcomes** | | | | | | | | | | | |
| Total score | 1 | 45 | 70.1 (19.1) | 39 | 71.2 (17.5) | 53 | 67.6 (19.7) | 53 | 71.8 (17.2) | 4.3 (0.9- 7.7) | 0.015 |
| | 2 | 45 | 72.8 (17.1) | 32 | 74.5 (15.7) | 48 | 72.0 (17.1) | 48 | 75.7 (15.5) | 3.7 (0.7- 6.8) | 0.017 |
| **Dimensions** | | | | | | | | | | | |
| Pain and hurt | 1 | 45 | 68.1 (27.1) | 39 | 73.4 (23.5) | 53 | 66.7 (27.0) | 53 | 73.8 (23.3) | 7.2 (-0.3- 14.7) | 0.060 |
| | 2 | 45 | 72.8 (26.0) | 32 | 75.4 (22.8) | 48 | 72.3 (26.2) | 48 | 74.7 (22.8) | 2.4 (-5.1- 10.0) | 0.51 |
| Nausea | 1 | 45 | 68.9 (27.4) | 39 | 74.5 (22.0) | 53 | 65.4 (28.7) | 53 | 73.9 (21.7) | 8.5 (3.5- 13.6) | 0.002 |
| | 2 | 45 | 73.4 (24.2) | 32 | 74.2 (23.7) | 48 | 71.6 (25.0) | 48 | 74.5 (23.9) | 2.9 (-1.4- 7.2) | 0.18 |
| Procedural anxiety | 1 | 45 | 68.3 (30.1) | 39 | 69.0 (29.4) | 53 | 68.7 (29.6) | 53 | 70.9 (28.2) | 2.1 (-6.1- 10.4) | 0.60 |
| | 2 | 45 | 73.5 (27.4) | 32 | 77.9 (27.2) | 48 | 74.8 (27.1) | 48 | 79.8 (25.9) | 5.0 (-1.0- 11.0) | 0.099 |
| Treatment anxiety | 1 | 45 | 81.9 (22.1) | 39 | 77.4 (26.1) | 53 | 79.6 (23.2) | 53 | 79.9 (24.5) | 0.3 (-5.5- 6.1) | 0.92 |
| | 2 | 45 | 82.6 (23.0) | 32 | 86.2 (24.1) | 48 | 83.0 (22.5) | 48 | 88.1 (22.2) | 5.1 (0.5- 9.8) | 0.031 |
| Worry | 1 | 45 | 70.9 (25.3) | 39 | 67.5 (25.5) | 53 | 67.6 (26.6) | 53 | 68.1 (24.9) | 0.5 (-5.5- 6.5) | 0.86 |
| | 2 | 45 | 72.4 (23.8) | 32 | 74.7 (21.8) | 48 | 71.5 (23.9) | 48 | 76.1 (21.7) | 4.6 (-0.6- 9.8) | 0.083 |
| Cognitive problems | 1 | 45 | 60.9 (25.2) | 39 | 62.7 (24.1) | 53 | 58.8 (24.9) | 53 | 61.8 (23.5) | 3.0 (-1.6- 7.6) | 0.20 |
| | 2 | 45 | 63.4 (20.9) | 32 | 61.1 (19.7) | 48 | 62.4 (21.6) | 48 | 63.5 (19.3) | 1.1 (-3.9- 6.0) | 0.66 |
| Perceived physical appearance | 1 | 45 | 69.8 (27.1) | 39 | 72.6 (26.5) | 53 | 68.1 (27.3) | 53 | 73.3 (26.0) | 5.2 (-1.9- 12.2) | 0.15 |
| | 2 | 45 | 73.9 (27.9) | 32 | 76.0 (30.6) | 48 | 72.3 (28.4) | 48 | 77.5 (30.0) | 5.2 (-2.0- 12.4) | 0.15 |
| Communication | 1 | 45 | 78.3 (20.8) | 39 | 76.5 (19.8) | 53 | 76.4 (21.6) | 53 | 76.8 (19.2) | 0.4 (-6.0- 6.7) | 0.91 |
| | 2 | 45 | 76.3 (19.5) | 32 | 79.4 (17.5) | 48 | 76.2 (19.7) | 48 | 78.9 (16.8) | 2.6 (-2.6- 7.9) | 0.32 |

N = Number; SD = Standard Deviation; Est = Estimated; CI = Confidence Interval

##### ii. Parent reported questionnaires

Parent-reported Quality of Life (QoL) was collected using Pediatric Quality of Life Inventory™ (PedsQL) for children aged 5-19 years. Higher scores indicate better health related QoL.

| | | Observed | | | | Imputed | | | | | |
|---|---|---|---|---|---|---|---|---|---|---|---|
| | | Pre | | Post | | Pre | | Post | | Difference | |
| | Dose | N | Mean (SD) | N | Mean (SD) | N | Mean (SD) | N | Mean (SD) | Est (95% CI) | p-value |
| **Primary outcomes** | | | | | | | | | | | |
| Total score | 1 | 54 | 72.1 (15.7) | 55 | 74.2 (16.7) | 70 | 71.5 (16.3) | 70 | 72.4 (16.8) | 0.9 (-2.0 - 3.7) | 0.55 |
| | 2 | 49 | 70.9 (17.8) | 45 | 73.1 (17.5) | 56 | 70.4 (17.4) | 56 | 73.5 (17.1) | 3.1 (0.1 - 6.1) | 0.046 |
| **Dimensions** | | | | | | | | | | | |
| Pain and hurt | 1 | 54 | 68.1 (25.4) | 55 | 75.9 (22.0) | 70 | 66.4 (26.1) | 70 | 73.9 (21.5) | 7.5 (1.8 - 13.1) | 0.011 |
| | 2 | 50 | 71.5 (24.7) | 46 | 72.6 (27.3) | 58 | 72.1 (24.7) | 58 | 72.5 (27.0) | 0.3 (-4.5 - 5.2) | 0.88 |
| Nausea | 1 | 54 | 73.1 (23.9) | 55 | 75.4 (24.3) | 70 | 72.5 (24.2) | 70 | 72.1 (25.0) | -0.4 (-5.4 - 4.7) | 0.88 |
| | 2 | 49 | 71.3 (25.1) | 46 | 74.2 (25.5) | 57 | 70.5 (24.8) | 57 | 74.0 (24.9) | 3.5 (-0.7 - 7.7) | 0.10 |
| Procedural anxiety | 1 | 54 | 71.1 (28.5) | 55 | 72.4 (28.1) | 70 | 72.6 (27.2) | 70 | 72.1 (27.9) | -0.5 (-6.1 - 5.1) | 0.86 |
| | 2 | 49 | 72.2 (29.5) | 45 | 74.4 (25.6) | 56 | 73.1 (28.3) | 56 | 75.0 (25.7) | 1.9 (-3.6 - 7.5) | 0.48 |
| Treatment anxiety | 1 | 54 | 81.2 (22.7) | 55 | 80.6 (26.5) | 70 | 79.5 (24.1) | 70 | 80.8 (25.9) | 1.3 (-4.3 - 6.8) | 0.65 |
| | 2 | 49 | 77.2 (27.6) | 45 | 80.9 (26.3) | 56 | 77.5 (26.7) | 56 | 82.0 (25.2) | 4.5 (-0.4 - 9.5) | 0.073 |
| Worry | 1 | 54 | 74.1 (21.6) | 55 | 75.0 (21.8) | 70 | 73.8 (21.5) | 70 | 74.1 (21.7) | 0.3 (-3.9 - 4.5) | 0.90 |
| | 2 | 49 | 71.3 (24.4) | 45 | 71.3 (23.6) | 56 | 69.5 (24.1) | 56 | 72.6 (23.7) | 3.1 (-1.5 - 7.7) | 0.18 |
| Cognitive problems | 1 | 53 | 60.9 (22.3) | 54 | 64.9 (21.4) | 70 | 60.3 (22.3) | 70 | 63.1 (21.5) | 2.8 (-1.7 - 7.4) | 0.22 |
| | 2 | 49 | 62.1 (21.0) | 45 | 63.3 (20.6) | 56 | 61.4 (21.2) | 56 | 63.9 (20.0) | 2.5 (-2.2 - 7.1) | 0.29 |
| Perceived physical appearance | 1 | 53 | 76.7 (24.4) | 54 | 75.8 (22.1) | 70 | 75.0 (24.5) | 70 | 74.7 (22.3) | -0.3 (-4.6 - 4.0) | 0.88 |
| | 2 | 49 | 70.9 (27.8) | 44 | 76.3 (26.4) | 56 | 70.5 (27.5) | 56 | 76.0 (25.9) | 5.4 (-2.0 - 12.8) | 0.15 |

| | | Observed | | | | Imputed | | | | | |
|---|---|---|---|---|---|---|---|---|---|---|---|
| | | Pre | | Post | | Pre | | Post | | Difference | |
| | Dose | N | Mean (SD) | N | Mean (SD) | N | Mean (SD) | N | Mean (SD) | Est (95% CI) | p-value |
| Communication | 1 | 52 | 75.7 (19.9) | 54 | 78.2 (20.8) | 70 | 76.1 (20.2) | 70 | 77.5 (20.8) | 1.4 (-3.1 - 6.0) | 0.52 |
| | 2 | 49 | 75.9 (20.7) | 44 | 78.0 (20.8) | 56 | 75.7 (20.8) | 56 | 77.7 (21.5) | 2.0 (-3.1 - 7.0) | 0.43 |

N = Number; SD = Standard Deviation; Est = Estimated; CI = Confidence Interval

**3n Baseline health-related quality of life and vaccine related beliefs**

Scores from the baseline Oxford vaccine Confidence and Complacency Scale were correlated with domains in the baseline quality of life questionnaires.

| QoL scale | N | Correlation (95% CI)* |
|---|---|---|
| **Adults (QLQ-C30)** | | |
| Emotional functioning | 246 | -0.18 (-0.31 - 0.04) |
| Global health status | 246 | -0.17 (-0.29 - 0.03) |
| Summary score | 241 | -0.17 (-0.30 - 0.04) |
| **Children** (PedsQL)** | | |
| Worry | 35 | -0.02 (-0.35 - 0.32) |
| Procedural anxiety | 35 | -0.13 (-0.44 - 0.21) |
| Treatment anxiety | 35 | -0.19 (-0.52 - 0.15) |
| Communication | 35 | -0.36 (-0.63 - 0.02) |
| Total score | 35 | -0.39 (-0.68 - 0.08) |

N = Number of responses; CI = Confidence Interval;
* using an estimated Spearman correlation coefficient r
** if child’s response was unavailable, parent response was used
